# Sporadic detection of dengue virus RNA in California wastewater solids

**DOI:** 10.64898/2026.10.26.26366224

**Authors:** Sheena Conforti, Mary E. Danforth, Andrea J. Lund, Hannah Romo, Stephanie Bertsch-Merbach, Alexander Yu, Alexandria B. Boehm

## Abstract

Dengue virus (DENV) is the most prevalent mosquito-borne virus globally, endemic in over 100 countries and associated with around 40,000 deaths each year. Prior research has shown that DENV RNA can be detected in wastewater, but detection rates vary widely depending on the setting and method used. In this study, we analyzed 1,072 untreated wastewater samples from five treatment plants in California, collected between December 2023 and December 2025, to evaluate wastewater-based surveillance for DENV. We compared wastewater data with reported clinical incidence and *Aedes aegypti* mosquito abundance measurements over the same period collected by the California Department of Public Health. DENV RNA was found in 1.2% of wastewater samples (13/1,072). DENV-1 was most common (0.6%, 6/1,072), followed by DENV-2 (0.4%, 4/1,072), and DENV-4 (0.3%, 3/1,072). No DENV-3 RNA was detected. DENV RNA was detected only in wastewater from southern California (plants in Los Angeles and San Diego counties), with no detections in wastewater from the Bay Area, including plants located in San Jose and Oakland. There was no correlation between wastewater DENV RNA levels and county-wide reported dengue incidence during the same week. However, counties with a higher incidence over the study period showed greater detection frequency of DENV RNA in wastewater. Weekly *Ae. aegypti* abundance was correlated with weekly incidence of locally-transmitted cases (Spearman’s ρ=0.45, p<0.01). Our results indicate that wastewater surveillance can detect DENV in areas of higher incidence, but the low detection frequency and sporadic nature of positive samples limit its utility as a standalone surveillance approach. Given the low incidence of dengue in California and the presence of robust clinical and mosquito surveillance systems, wastewater monitoring does not currently add significant public health value for dengue surveillance in the state.

## 1 Introduction

Dengue virus (DENV) is an enveloped arbovirus that comprises four antigenically distinct serotypes (DENV-1 to DENV-4) (1). DENV is the most prevalent mosquito-borne viral infection worldwide, is endemic in more than 100 countries and causes an estimated 40,000 deaths annually (2,3). Symptomatic infection typically presents with high fever, severe myalgia, and arthralgia lasting up to seven days (4). Over recent decades, reported dengue cases have increased substantially, roughly 30-fold between 2000 and 2024, making it the fastest-growing viral disease globally (2).

Transmission occurs primarily through bites of infected female *Aedes* mosquitoes (5), predominantly *Ae. aegypti* and *Ae. albopictus* (6,7). Globally, DENV spread is driven by international travel and trade, warming climates, rapid urbanization, asymptomatic infections, and vector adaptation, complicated by poor or inadequate surveillance (8–10). For example, *Ae. aegypti* had not been described in California until 2013, but is now well established in 28 California counties. In 2023 and 2024, 20 locally acquired dengue cases were documented across Los Angeles, San Diego, and San Bernardino counties (11), the first cases ever reported in California.

Previous studies have demonstrated the feasibility of detecting DENV RNA in wastewater, although reported detection frequencies vary substantially across settings and analytical approaches. DENV RNA has been detected in approximately 21% of wastewater samples during local transmission in Florida, 25% of wastewater samples in Portugal, one-third of wastewater samples in Italy, more than 90% of wastewater samples in Brazil, whereas only 2% of wastewater samples were positive in China (12–17). Of these, several studies that examined the relationship between wastewater DENV detections and clinical case counts found no clear correspondence, including a recent study from Brazil that reported no correlation between wastewater DENV RNA concentrations and case counts despite frequent detections (13–15,17). Together, these findings demonstrate that wastewater detection frequencies vary considerably across settings, and that where clinical comparisons have been made, wastewater DENV detections have not consistently corresponded with reported case counts, highlighting an incomplete understanding of the factors governing DENV RNA occurrence in wastewater.

One component of dengue surveillance is entomological surveillance, including monitoring *Aedes* mosquito populations and, in some cases, testing field-collected mosquitos for DENV RNA (11). DENV has occasionally been detected in mosquito pools before or without reported human cases in highly endemic settings, but mosquito pool testing is not highly sensitive and despite testing thousands of pools, many of which were in response to locally-acquired cases, only one *Ae. aegypti* pool has tested positive in California (18,19). Because mosquito abundance and wastewater-based surveillance capture different aspects of dengue epidemiology, evaluating them together may provide a more comprehensive understanding of DENV incidence and help interpret wastewater detections relative to reported dengue cases.

In this study, we evaluated wastewater-based surveillance for DENV by quantifying DENV RNA in 1,072 samples from five wastewater treatment plants (WWTPs) in California collected over time periods spanning December 2023 to December 2025. To contextualize these results, we integrated wastewater measurements with clinical case counts and mosquito abundance data collected over the same time period. This integrated approach allowed us to evaluate whether wastewater surveillance can complement existing dengue monitoring systems and inform public health responses in low-incidence settings.

## 2 Materials and methods

### 2.1 Wastewater samples collection

Wastewater samples were collected from five WWTPs in California, United States (**Figure 1**). Sites included the San Francisco Bay Area (San Jose, serving population = 1,500,000; Oakland, 740,000), and Southern California (Los Angeles - Carson, 3,500,000; Los Angeles - Playa del Rey, 4,000,000; San Diego, 2,200,000). Sites were selected for their relevance to dengue incidence. The Southern California sites were included because *Ae. aegypti* is well established in these counties and locally acquired dengue cases have been documented there in recent years (11). The Bay Area sites (San Jose, Oakland) were included to evaluate whether DENV RNA could be detected in wastewater in areas with lower overall travel-associated dengue incidence, a region without an established *Ae. aegypti* population, and no documented local transmission. At each site, 50 mL of 24-hour composite raw influent was collected in sterile containers three times per week. Sampling periods varied by site from December 2023 to December 2025 (Table 1). Samples were shipped at 4°C and processed within 1-3 days. Wastewater solids were isolated from the influent by settling for 15 minutes, and the settled fraction was aspirated into a separate tube for downstream processing. A total of 1,072 samples were analyzed.

**Table 1.** Sampling coverage and dengue virus (DENV) serotype RNA detection metrics across wastewater treatment plants (WWTP) included in the study. For each WWTP, the table reports the sampling period (first to last collection date) and the total number of wastewater samples analysed (n). “Any DENV RNA detected” indicates samples in which at least one of the four serotype-specific RT-ddPCR assays (DENV-1 to DENV-4) yielded a quantified concentration. For each dengue virus serotype (DENV-1 to DENV-4), the number of samples in which RNA of the serotype was detected and the corresponding detection prevalence are shown as n (%). Detection was defined as a quantified DENV RNA concentration above the assay detection threshold in the corresponding serotype-specific RT-ddPCR assay (e.g., values not reported as “ND”). Detection prevalence (%) was calculated as the proportion of samples with detection of the respective serotype divided by the total number of samples analysed for that plant. The first row (“All WWTPs”) reports the same metrics pooled across all WWTP.

| WWTP | Sampling period | Total samples (n) | Any DENV detected (n, %) | DENV-1 detected (n, %) | DENV-2 detected (n, %) | DENV-3 detected (n, %) | DENV-4 detected (n, %) |
| --- | --- | --- | --- | --- | --- | --- | --- |
| All WWTPs | 2023-12-17 to 2025-12-31 | 1072 | 13 (1.2%) | 6 (0.6%) | 4 (0.4%) | 0 (0.0%) | 3 (0.3%) |
| Los Angeles - Carson, CA | 2023-12-17 to 2025-12-29 | 317 | 7 (2.2%) | 4 (1.3%) | 2 (0.6%) | 0 (0.0%) | 1 (0.3%) |
| Los Angeles - Playa del Rey, CA | 2024-06-23 to 2025-12-29 | 232 | 4 (1.7%) | 2 (0.9%) | 1 (0.4%) | 0 (0.0%) | 1 (0.4%) |
| Oakland, CA | 2023-12-19 to 2024-06-20 | 81 | 0 (0.0%) | 0 (0.0%) | 0 (0.0%) | 0 (0.0%) | 0 (0.0%) |
| San Diego, CA | 2024-06-23 to 2025-12-29 | 213 | 2 (0.9%) | 0 (0.0%) | 1 (0.5%) | 0 (0.0%) | 1 (0.5%) |
| San Jose, CA | 2024-07-21 to 2025-12-31 | 229 | 0 (0.0%) | 0 (0.0%) | 0 (0.0%) | 0 (0.0%) | 0 (0.0%) |

**Figure 1.**
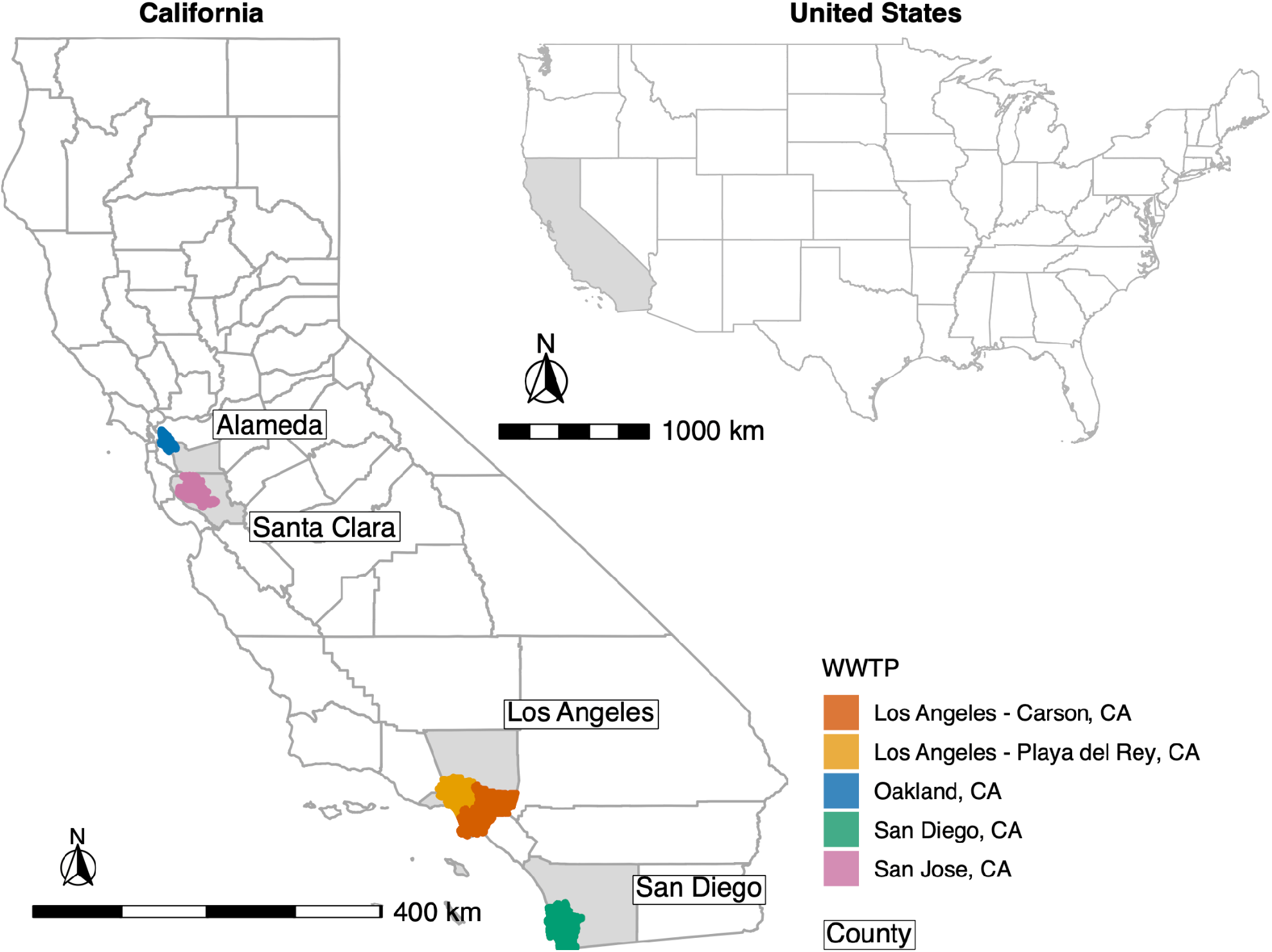
Geographic distribution of wastewater treatment plant (WWTP) sewersheds included in this study. The map shows the selected California WWTP sewershed boundaries, colored by WWTP, with the corresponding counties highlighted in light grey. The inset shows the location of California within the contiguous United States. Scale bars and north arrows are shown for geographic reference.

### 2.2 Wastewater samples preanalytical processing and RNA extraction

Preanalytical processing and RNA extraction were performed as previously described (17). Briefly, wastewater solids were dewatered by centrifugation and resuspended in DNA/RNA Shield (Zymo Research, Irvine, CA) at 0.75 mg wet weight/mL. Dry weight was determined from a separate aliquot of dewatered solids. RNA was extracted from six replicate aliquots (ten replicate aliquots for the WWTP San Jose) using the Chemagic Viral DNA/RNA 300 Kit H96 on the PerkinElmer Chemagic 360 platform (PerkinElmer, Waltham, MA). PCR inhibitors were then removed with the Zymo OneStep-96 PCR Inhibitor Removal Kit (Zymo Research). A 300 µL aliquot of the suspension was processed, and nucleic acids were eluted in 60 µL (20).

### 2.3 Quantification of DENV RNA in wastewater samples

Digital droplet reverse transcription PCR (RT-ddPCR) was used to quantify DENV RNA in wastewater samples as previously described (17). DENV serotypes 1 to 4 were quantified using previously published serotype-specific multiplex assays (Supplementary Table 1) (21). RNA extracts were analyzed in six replicates per sample (ten replicates for San Jose, Santa Clara County) using a Bio-Rad AutoDG system, thermal cycling, and droplet reading on QX200 or QX600 platforms (Supplementary Data 1.1). For DENV-3, a degenerate probe was implemented from February 18, 2025 onwards, replacing the original probe to account for sequence variability. Assay specificity and sequence coverage were evaluated *in silico* against publicly available dengue virus genomes and experimentally validated for selected variants (Supplementary Data 1.2, Supplementary Table 2). Each 96-well PCR plate included no-template PCR controls and positive controls. Positive controls consisted of ATCC dengue virus reference genomic RNA for each serotype (Serotype 1: VR-3228SD; Serotype 2: VR-3229SD; Serotype 3: VR-3230SD; Serotype 4: VR-3231SD). Samples were considered positive if ≥3 positive droplets were detected across merged replicates. Concentrations were reported as copies per gram dry weight (cp/g).

### 2.4 Mosquito surveillance data

Mosquito surveillance data, originating from specimens collected by local vector control agencies, are stored in an online data management system, the California Vectorborne Disease Surveillance Gateway (VectorSurv, https://vectorsurv.org). Data for this project were obtained via data request #000097 for Los Angeles, San Diego, Alameda, and Santa Clara counties for the period of December 2023 to December 2025. Data were filtered to remove entries associated with trap malfunctions. Species names were standardized, and analyses were restricted to *Ae. aegypti*, the primary vector of dengue virus globally (2). Data collected from multiple trap types were retained to maximize spatial and temporal coverage, with BG Sentinel traps comprising the majority of *Ae. aegypti* collections (Supplementary Figure 2). Trap collection frequency is determined independently by each vector control agency and is not mandated or standardized across jurisdictions; the temporal resolution of the dataset can be as fine as daily, though collection intervals vary by site, trap type, and season. For each trap collection event, the dataset reports the number of mosquitoes counted, the number of traps deployed, and the number of nights each trap was active. Raw counts were converted to mosquito abundance by dividing the number of *Ae. aegypti* collected by the number of trap-nights, where one trap-night represents a single trap deployed for one night. Mosquito abundance was calculated daily for each county and summarized as a 15-day rolling average to characterize temporal trends over the study period.

**Figure 2.**
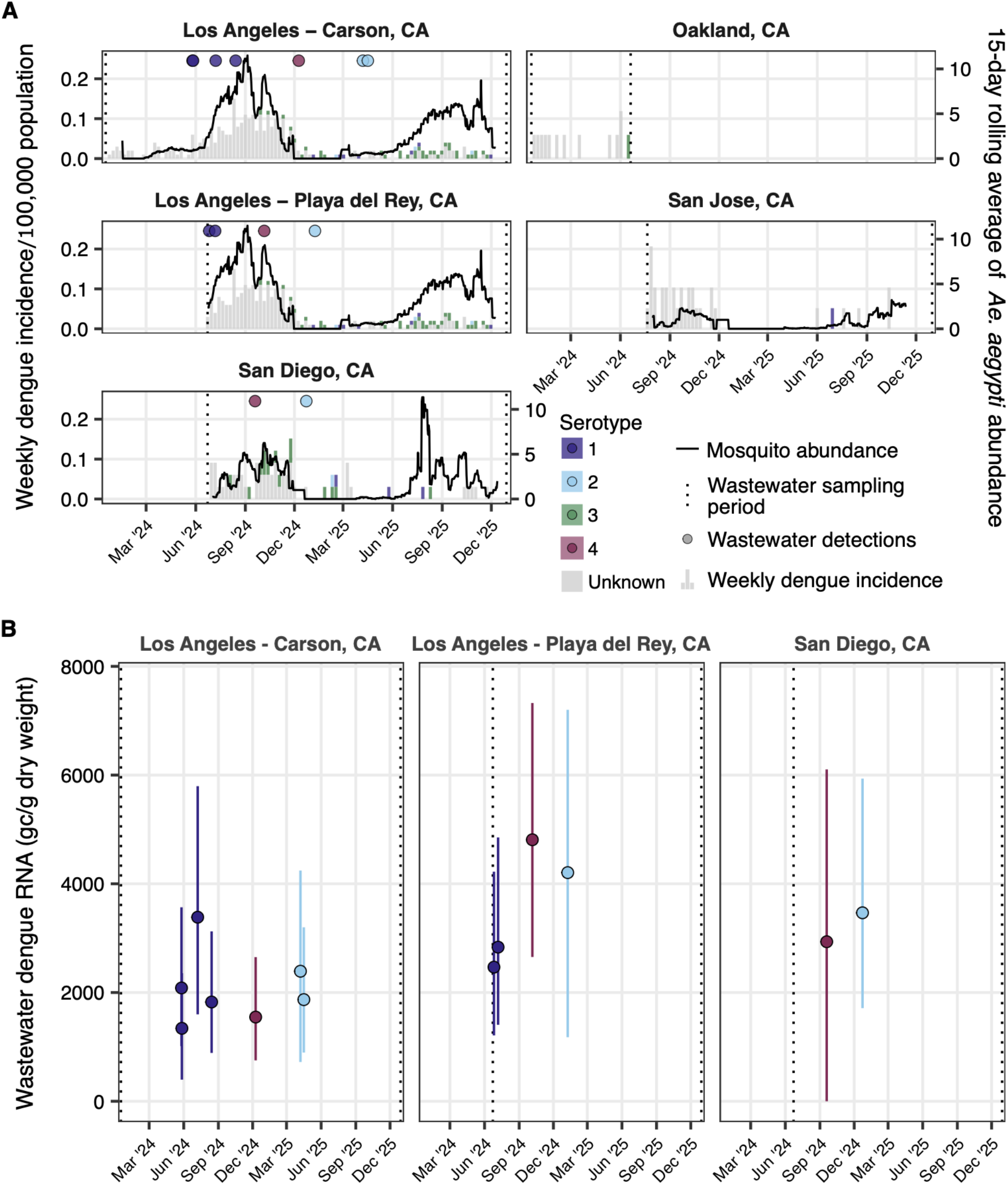
Longitudinal comparison of county-level reported dengue incidence, *Aedes aegypti* abundance, and wastewater dengue virus (DENV) detections and measured RNA concentrations across California wastewater treatment plants (WWTPs) from December 2023 to December 2025. (A) Weekly reported dengue incidence (per 100,000 population) aggregated at the county level and stratified by serotype are shown as bars. Colored circles indicate dates of wastewater DENV RNA detections, with colors denoting DENV serotype (1-4). The black line represents the centered 15-day rolling average of *Ae. aegypti* abundance, standardized by trapping effort (mosquitoes per trap-night), shown on the right y-axis. Vertical dotted lines indicate the start and end of the wastewater sampling period for each site. Mosquito abundance data were visualized only for sites located in counties with *Ae. aegypti* detections (there were no detections in Alameda County where the Oakland, CA site is located) during the wastewater sampling period. (B) Wastewater DENV RNA concentrations (gc/g dry weight) quantified by serotype-specific RT-ddPCR assays, shown only for the subset of samples in which DENV was detected; non-detects are excluded from this panel. Points represent measured concentrations among these DENV-positive samples and vertical error bars indicate 95% confidence intervals. Colors denote DENV serotype (1-4). Vertical dotted lines indicate the wastewater sampling period for each site. Only WWTPs with detectable DENV RNA are shown.

### 2.5 Reported dengue cases in California

Reported dengue case data for Alameda, Santa Clara, Los Angeles, and San Diego counties, the four counties containing the investigated WWTPs, were obtained from the California Department of Public Health (CDPH). These data are collected as part of routine public health surveillance and were therefore exempt from Institutional Review Board (IRB) review and approval at the state level; the overall study was conducted under IRB approval from Stanford University (protocol 86402). The dataset covered the period from December 2023 to December 2025 and included symptom onset date, reported serotype, when available, and whether the case was classified as travel-associated or locally-acquired. Cases were aggregated by county at weekly and monthly resolution based on symptom onset date to enable comparison with wastewater dengue virus detections and *Ae. aegypti* abundance. Dengue incidence was calculated as reported cases divided by county population (22), multiplied by 100,000.

### 2.6 Data merging and statistical analysis

Statistical analyses were performed in R (v4.4.1) and RStudio (v2024.04.2). Wastewater data were linked to the mosquito abundance and clinical case datasets by county. Mosquito and human case reports were then restricted temporally to the wastewater sampling window for each county, defined by the earliest and latest wastewater collection dates pooled across the WWTPs in each county.

Wastewater signal was compared to reported dengue incidence and *Ae. aegypti* abundance at three resolutions. At the sample level, DENV concentration in each wastewater sample (summed across serotypes; non-detects set to 0) was compared to reported dengue incidence county-wide during the corresponding week. At the county level, the proportion of DENV-positive samples pooled across WWTPs over the full sampling period was compared descriptively (n = 4 counties) to cumulative reported dengue incidence over the same period. Cumulative dengue incidence is defined at the total number of reported cases over the study period dived by the total population of the county and multiplied by 100,000. Weekly *Ae. aegypti* abundance was compared to weekly reported dengue incidence, considering all cases and locally acquired cases separately. Case counts were aggregated to Sunday-starting epidemiological weeks, and weeks without a reported case within the sampling window of the corresponding county were retained as zero counts.

Associations were assessed using Spearman’s rank correlation (rho); p-values are reported for the sample- and week-level comparisons, but not for the county-level comparison, where n = 4 leaves any significance test uninformative.

## 3 Results

We analyzed 1,072 wastewater samples collected from five WWTPs in California between December 2023 and December 2025 using serotype-specific RT-ddPCR assays targeting dengue virus (DENV) serotypes 1-4 RNA (Table 1). Out of the 856 control reactions run (428 no-template and 428 ddRT-PCR positive control reactions), all no-template controls were negative and all ddPCR positive controls were positive (Supplementary Table 3). DENV RNA was detected in 1.2% (n = 13) of all samples. Among serotypes, DENV-1 RNA was most frequently detected (0.6%, n = 6), followed by DENV-2 (0.4%, n = 4), and DENV-4 (0.3%, n = 3). DENV-3 RNA was not detected in any of the samples.

Detection prevalence varied across WWTPs. DENV RNA was detected in wastewater from Southern California WWTPs, namely Los Angeles - Carson, Los Angeles - Playa del Rey, and San Diego, but not in Bay Area WWTPs, namely Oakland and San Jose (Figure 2). The highest proportion of DENV-positive samples was observed in Los Angeles - Carson (2.2%, n = 7/317). DENV RNA was also detected in 1.7% (n = 4/232) of samples from Los Angeles - Playa del Rey and 0.9% (n = 2/213) from San Diego. At Los Angeles - Carson, DENV-1 RNA was detected in 1.3% (n = 4/317) of samples, DENV-2 in 0.6% (n = 2/317), and DENV-4 in 0.3% (n = 1/317) (Table 1). At Los Angeles - Playa del Rey, DENV-1, DENV-2, and DENV-4 RNA were each detected in 0.4% (n = 1/232) of samples. In San Diego, DENV-2 and DENV-4 RNA were each detected in 0.5% (n = 1/213) of samples. DENV-3 RNA was not detected during the study period. No sample contained RNA from more than one serotype. Among samples with detectable DENV RNA, concentration distributions were broadly comparable across serotypes and WWTPs, with overlapping ranges and similar central tendencies (Supplementary Figure 1, Supplementary Table 4 and S5).

Mosquito abundance differed significantly among counties (Kruskal-Wallis, *p* < 0.001; Supplementary Table 6, Supplementary Figure 3). Los Angeles County exhibited the highest county-level daily *Ae. aegypti* abundance (mean ± standard deviation (SD): 1.6 ± 3.2), followed by San Diego (1.4 ± 7.7) and Santa Clara County (0.2 ± 0.7), whereas no *Ae. aegypti* mosquitoes were detected in Alameda County during the study period (Supplementary Table 7). Pairwise comparisons confirmed higher mosquito abundance in Los Angeles than in San Diego (p-adj = 0.001) and Santa Clara (p-adj < 0.001), as well as higher abundance in San Diego than in Santa Clara (p-adj < 0.001) (Supplementary Table 6). Mosquito populations displayed marked seasonality, with abundance increasing during summer and autumn and declining during winter months in both 2024 and 2025 (Supplementary Figure 3). Within counties, daily mosquito abundance did not differ significantly between 2024 and 2025 in any of the counties (Supplementary Table 8).

**Figure 3.**
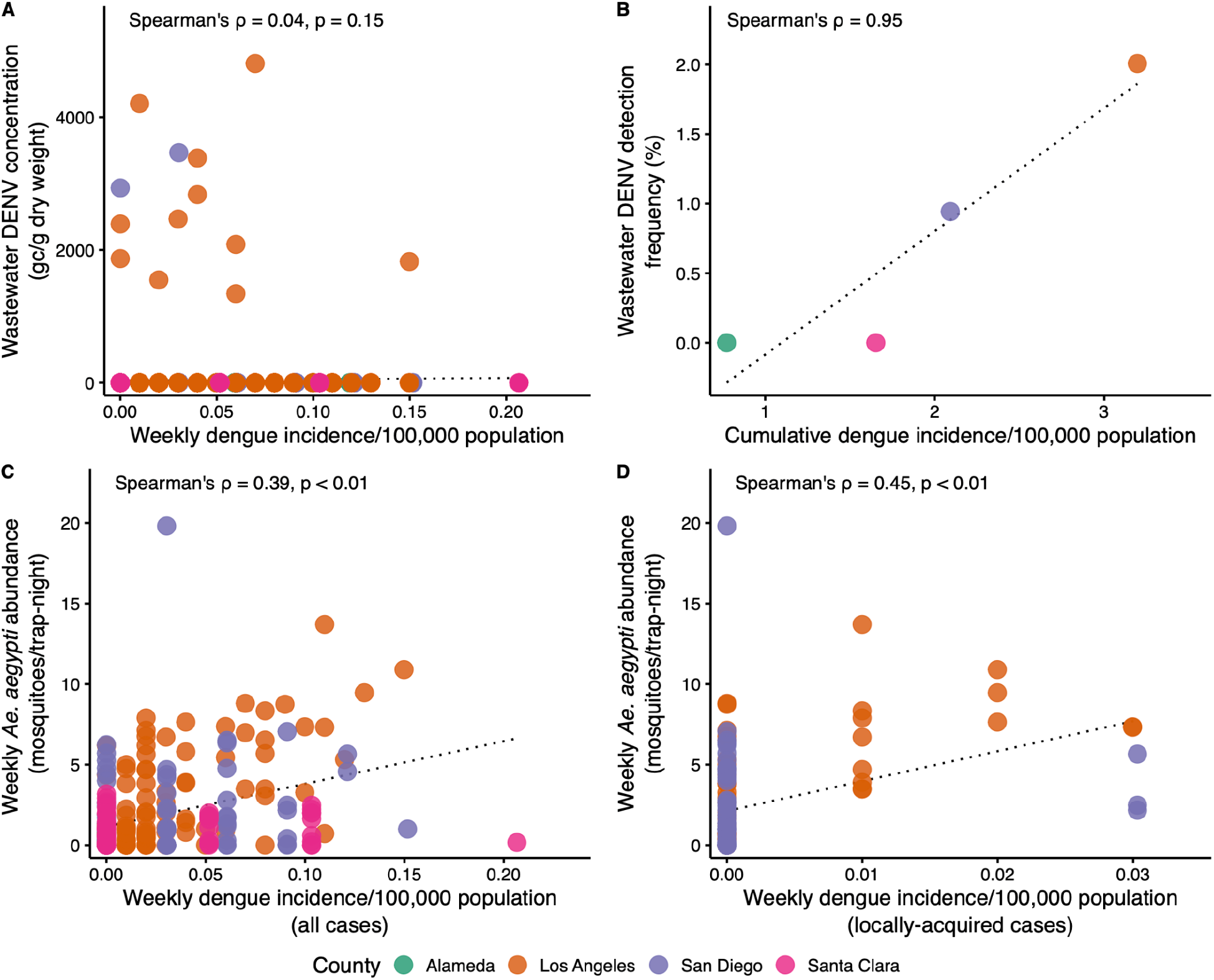
Comparison of wastewater dengue virus (DENV) RNA signal, *Aedes aegypti* abundance, and dengue incidence across California sites, colored by county. (A) Wastewater DENV RNA concentration (gc/g dry weight; non-detects set to 0) for each sample versus the dengue incidence county-wide in the same week of sample collection. (B) Wastewater DENV RNA detection frequency versus the cumulative dengue incidence over the whole study period, one point per county. (C) Weekly *Ae. aegypti* abundance versus weekly dengue incidence, one point per county-week. (D) Same as (C), restricted to locally acquired cases only. The black dotted line in each panel is a linear trend line with Spearman’s ρ (with p-values in panels A, C, D; omitted in panel B, where n = 4 counties leaves both tests powerless) annotated (p-values below 0.005 are shown as p < 0.01).

Dengue cases were reported in all four counties during the study period, although incidence varied substantially by location (Figure 2, Supplementary Table 9). Los Angeles County reported the highest cumulative dengue incidence over the study period (3.2 cases/100,000 population), followed by San Diego (2.1 cases/100,000 population), Santa Clara (1.7 cases/100,000 population), and Alameda (0.8 cases/100,000). Dengue incidence peaked during summer and autumn 2024, particularly in Los Angeles County (Supplementary Figure 4A). San Diego exhibited a similar but less pronounced temporal pattern, whereas Santa Clara and Alameda reported comparatively a lower incidence and only sporadic wastewater detections. Most reported cases lacked serotype information, accounting for 79-97% of cases in each county. Among typed cases, DENV-3 was the most frequently reported serotype, particularly in Los Angeles (12% of all cases) and San Diego (16%), whereas DENV-4 was not reported in any county (Supplementary Table 9). The majority of cases in each county were travel-associated, ranging from 94% in Los Angeles to 100% in Santa Clara and Alameda. Locally acquired cases were reported only in Los Angeles (n = 20/320, 6%) and San Diego (n = 3/69, 4%) (Supplementary Table 9, Supplementary Figure 4B).

To assess whether wastewater surveillance reflected independent measures of dengue activity, we compared wastewater DENV RNA signal, *Ae. aegypti* abundance, and reported dengue incidence at the sample, county, and week level. At the sample level, wastewater DENV RNA concentration did not correlate with the dengue incidence reported county-wide during the same week (Spearman’s ρ = 0.04, p = 0.15; Figure 3A). At the county level, the proportion of DENV RNA-positive wastewater samples over the full sampling period was strongly correlated with the cumulative dengue incidence per county (Spearman’s ρ = 0.95; Figure 3B, Supplementary Table 10). Counties with the highest dengue incidence had the highest wastewater detection frequency: Los Angeles ranked highest on both, San Diego second, and Alameda and Santa Clara lowest, with no wastewater detections at all. Weekly *Ae. aegypti* abundance was positively correlated with weekly dengue incidence, both considering all cases (Spearman’s ρ = 0.39, p < 0.01; Figure 3C) and when restricted to locally acquired cases (Spearman’s ρ = 0.45, p < 0.01; Figure 3D).

## 4 Discussion

In this study, we evaluated wastewater-based surveillance for DENV in California by comparing wastewater measurements with human case data and mosquito surveillance. DENV RNA was detected in wastewater from three WWTPs in Southern California (two in Los Angeles and one in San Diego). However, detections were sporadic, occurring in only 1.2% (13/1,072) of samples collected over two years. DENV RNA was not detected in wastewater from the Bay Area of California, neither in Oakland nor San Jose. Together, these findings demonstrate that DENV can be detected in municipal wastewater but suggest that sensitivity of wastewater surveillance is limited in settings where incidence is low, as in California.

Several biological and methodological factors may explain the low detection frequency observed. Unlike enteric viruses, DENV is a blood-borne, mosquito-transmitted pathogen and does not extensively replicate in the gastrointestinal tract. Consequently, viral RNA is expected to enter wastewater at relatively low concentrations through urine, saliva, and other bodily fluids rather than sustained fecal shedding. Previous studies have shown that DENV RNA can be detected in urine and saliva and shedding may persist for several weeks following initial infection (5,23). Nevertheless, once released into sewer systems, viral RNA is subject to dilution and potentially environmental degradation (24), and detection may be further hindered by wastewater constituents such as organic matter, detergents, and PCR inhibitors (5). In our study, we took a number of steps to overcome potential inhibition including using digital PCR, inhibitor removal step, and dilution of solids in the buffer at a level thoughtfully titrated to alleviate inhibition (25). Overall, limited shedding of DENV RNA probably limits our ability to detect DENV RNA in this setting, where Los Angeles County, our highest incidence site, had a cumulative incidence of only 3.2 reported cases per 100,000 population over the full two-year study period (320 cases total). Interestingly, incidence rates from locations where DENV RNA was more prevalent in wastewater were three orders of magnitude higher than those observed here. In Belo Horizonte, Brazil, confirmed dengue cases reached nearly 200,000 in 2024 alone, in a metropolitan area of about 2.7 million people, corresponding to an incidence of roughly 7,400 cases per 100,000 population in that single year (15). DENV RNA might only be consistently detectable in wastewater at higher incidence rates like those observed in Brazil.

Serotype composition differed between reported human cases and wastewater detections. Most human cases had no reported serotype since laboratory diagnosis is often determined by serological rather than molecular testing. Among the subset with a reported serotype, DENV-3 was the most common. In wastewater, however, DENV-3 was never detected, a finding that also differs from Wolfe et al., who detected only DENV-3 in wastewater from Florida (17). This discrepancy may reflect serotype-specific differences in shedding or assay sensitivity rather than an absence of DENV-3 in wastewater. Conversely, DENV-4, of which RNA was detected in wastewater but never reported among typed human cases, may be disproportionately asymptomatic or otherwise underdiagnosed, though this remains speculative given the small number of detections.

The geographic distribution of wastewater detections mirrored differences in mosquito abundance and dengue incidence across four California counties. Wastewater DENV RNA detections occurred exclusively in Southern California, where *Ae. aegypti* populations were established and dengue incidence was higher. In contrast, no wastewater detections were observed in Alameda County, where *Ae. aegypti* mosquitoes were absent during the study period, or in Santa Clara County, where mosquito abundance and dengue incidence remained low. The relationship observed between wastewater DENV detection, *Ae. aegypti* abundance, and dengue incidence are correlational and should not be interpreted as evidence of a causal relationship. The majority of reported cases in our study counties were travel-associated, rather than locally acquired, thus local *Ae. aegypti* populations were not the direct cause of most infections. The associations at the county and weekly levels more plausibly reflect shared seasonal and geographic patterns, such as increased *Ae. aegypti* abundance, travel volume, and case reporting occurring concurrently during summer and autumn in counties where the vector is established, rather than a direct mechanistic relationship between local mosquito populations and imported cases.

This study has several limitations. Human case counts reflect reported infections and likely underestimate true dengue incidence, given frequent asymptomatic or mild infections and potential underdiagnosis of travel-associated cases. Mosquito abundance data were collected independently by multiple vector control agencies using non-standardized trap types and collection frequencies, which may introduce heterogeneity across counties beyond true differences in abundance. Sampling periods also differed substantially across WWTPs (e.g., roughly six months at Oakland versus two years in Los Angeles - Carson), so sites with shorter or less representative windows had fewer opportunities to detect DENV in wastewater, limiting direct comparability across locations. Reassuringly, assay specificity and sensitivity to circulating sequence variants were evaluated both *in silico* and experimentally, with detection retained across all tested variants for every serotype. However, the DENV-3 probe was updated through the study to better accommodate observed sequence variability, so a modest degree of differential DENV-3 sensitivity before versus after that update cannot be fully excluded.

Taken together, our results indicate that DENV RNA can be detected in wastewater in areas experiencing a higher dengue incidence, but the low detection frequency and sporadic nature of positive samples limit its utility as a standalone surveillance approach in California. Reliable wastewater detection may require community-level dengue incidence well above that observed in California during this study period. Future studies should evaluate whether next-generation sequencing methods can improve detection sensitivity and serotype characterization. Established environmental surveillance approaches targeting mosquito breeding habitats, such as standing water bodies or stormwater systems, may also prove more informative for monitoring mosquito-borne pathogens than municipal wastewater. Given the low incidence of dengue in California and the presence of robust public health and mosquito surveillance, wastewater monitoring did not add significant public health value for dengue surveillance in the state during this study period. Dengue surveillance will likely continue to rely primarily on clinical and entomological monitoring, with wastewater serving as a complementary source of information rather than a primary surveillance tool, particularly in low-incidence settings such as California.

## 6 Author Contributions

S.C.: conceptualization, data curation, formal analysis, investigation, methodology, validation, visualization, writing - original draft, and writing - review & editing. M.E.D.: data curation, writing - review & editing. A.J.L.: data curation, writing - review & editing. H.R.: data curation, writing - review & editing. S.B.M.: writing - review & editing. A.Y.: conceptualization, writing - review & editing. A.B.B.: conceptualization, funding acquisition, project administration, resources, supervision, validation, writing - review & editing.

## 7 Data Availability Statement

All data used in this study, along with the R code for statistical analyses, are publicly available at the Stanford Digital Repository: https://purl.stanford.edu/xv431zh7260.

## 9 Acknowledgments

We thank the participating wastewater treatment plants for their samples for the project.

