## Supplementary material for "Sporadic detection of dengue virus RNA in California wastewater solids"

### 1 Supplementary Data

#### 1.1 ddPCR reaction setup and thermocycling conditions

Digital droplet RT-PCR (RT-ddPCR) assays were performed using a multiplex approach to simultaneously quantify DENV-1, DENV-2, DENV-3, and DENV-4 within the same reaction. Samples were analyzed on 96-well plates, with nine wastewater samples per plate for the DENV multiplex assay, each processed in ten replicate wells. Each plate also included three replicate negative extraction controls (buffer), two replicate no-template PCR controls (molecular-grade water), and one positive control (cDNA). For pepper mild mottle virus (PMMoV) and bovine coronavirus (BCoV), two replicate wells per sample were analyzed in duplex, as previously described (1).

RT-ddPCR reactions were prepared in a total volume of 22 µL, of which 20 µL was partitioned into droplets. Each reaction contained 5.5 µL of RNA template, 5.5 µL of One-Step RT-ddPCR Advanced Kit for Probes (Bio-Rad), 2.2 µL of reverse transcriptase (200 U/µL), 1.1 µL of 300 mM dithiothreitol (DTT), and primers and probes at final concentrations of 900 nM and 250 nM, respectively. Primers and probes were obtained from Integrated DNA Technologies (IDT, San Diego, CA) (Supplementary Table 1). The DENV multiplex assay was performed on undiluted RNA template, whereas PMMoV and BCoV assays were conducted in duplex on template diluted 1:100 in molecular-grade water.

Droplets were generated using the Bio-Rad AutoDG Automated Droplet Generator. Thermal cycling was performed on a Mastercycler Pro (Eppendorf) under the following conditions: reverse transcription at 50°C for 60 minutes, enzyme activation at 95°C for 5 minutes, followed by 40 cycles of denaturation at 95°C for 30 seconds and annealing/extension at 59°C for 30 seconds for the DENV multiplex assay, or 56°C for PMMoV/BCoV assays. A final enzyme deactivation step was performed at 98°C for 10 minutes, followed by a hold at 4°C for at least 30 minutes to stabilize droplets. The ramp rate was set to 2°C per second.

Droplet fluorescence was measured using either a QX200 or QX600 Droplet Reader (Bio-Rad). Wells containing fewer than 10,000 accepted droplets were excluded from analysis. Data were processed using QuantaSoft Analysis Pro (Bio-Rad) and QX Manager software, and replicate wells were merged prior to quantification. A sample was considered positive if at least three positive droplets were detected across merged wells.

#### 1.2 *In silico* assay screening

Primer and probe sequences for the serotype-specific DENV RT-ddPCR assays were screened *in silico* against publicly available dengue virus genomes downloaded from NCBI using a custom

Python script (available at https://purl.stanford.edu/xv431zh7260). Analyses were restricted to human-derived sequences with reported collection dates between July 1, 2023 and December 31, 2025. Sequences lacking collection dates were excluded. Sequences were classified as "pass" if the best candidate amplicon met the predefined mismatch thresholds of ≤2 mismatches in each primer and ≤1 mismatch in the probe. Sequences were classified as "fail" if no candidate amplicon satisfied these thresholds. "Pass" sequences were further stratified based on mismatch profiles into (i) perfect matches (no mismatches), (ii) primer mismatches only, and (iii) probe mismatches (Supplementary Table 2).

For DENV-1, 41.3% of sequences showed perfect matches, while 50.1% contained primer mismatches only, 5.3% probe mismatches, and 3.3% failed to meet screening thresholds (Supplementary Table 2). For DENV-2, only 10.0% of sequences showed perfect matches, with the majority exhibiting probe mismatches (59.7%), followed by failed screens (22.6%) and primer mismatches only (7.7%). For DENV-3, the proportion of perfectly matched sequences depended on probe configuration, with 83.5% perfect matches for DENV-3 degenerated probe compared to 16.0% for DENV-3 original, and correspondingly lower probe mismatch rates (10.1% vs 78.6%). For DENV-4, 70.1% of sequences showed perfect matches, with smaller proportions exhibiting primer mismatches only (9.2%), probe mismatches (14.9%), or failing screening (5.7%).

To further evaluate assay performance under conditions of observed sequence variation, experimental validation was conducted for DENV-1 and DENV-2 using synthetic gene blocks representing circulating variants containing single nucleotide polymorphisms (SNPs) within primer and probe binding regions. These serotypes were selected based on the *in silico* screening results, which showed lower proportions of perfectly matched sequences (41.3% for DENV-1 and 10.0% for DENV-2) and higher frequencies of mismatches compared to DENV-3 (83.5%) and DENV-4 (70.1%). For both DENV-1 and DENV-2, assays detected variant sequences at an input concentration of 25 copies per reaction, with measured concentrations ranging from 0.89 to 1.22 copies/µL for DENV-1 and 0.98 to 1.67 copies/µL for DENV-2, depending on the variant tested. Minor reductions in measured concentrations were observed for some variants relative to reference controls, indicating modest effects of sequence mismatches on assay efficiency; however, detection was consistently retained across all evaluated variants.

### 2 Supplementary Figures and Tables

#### 2.1 Supplementary Figures

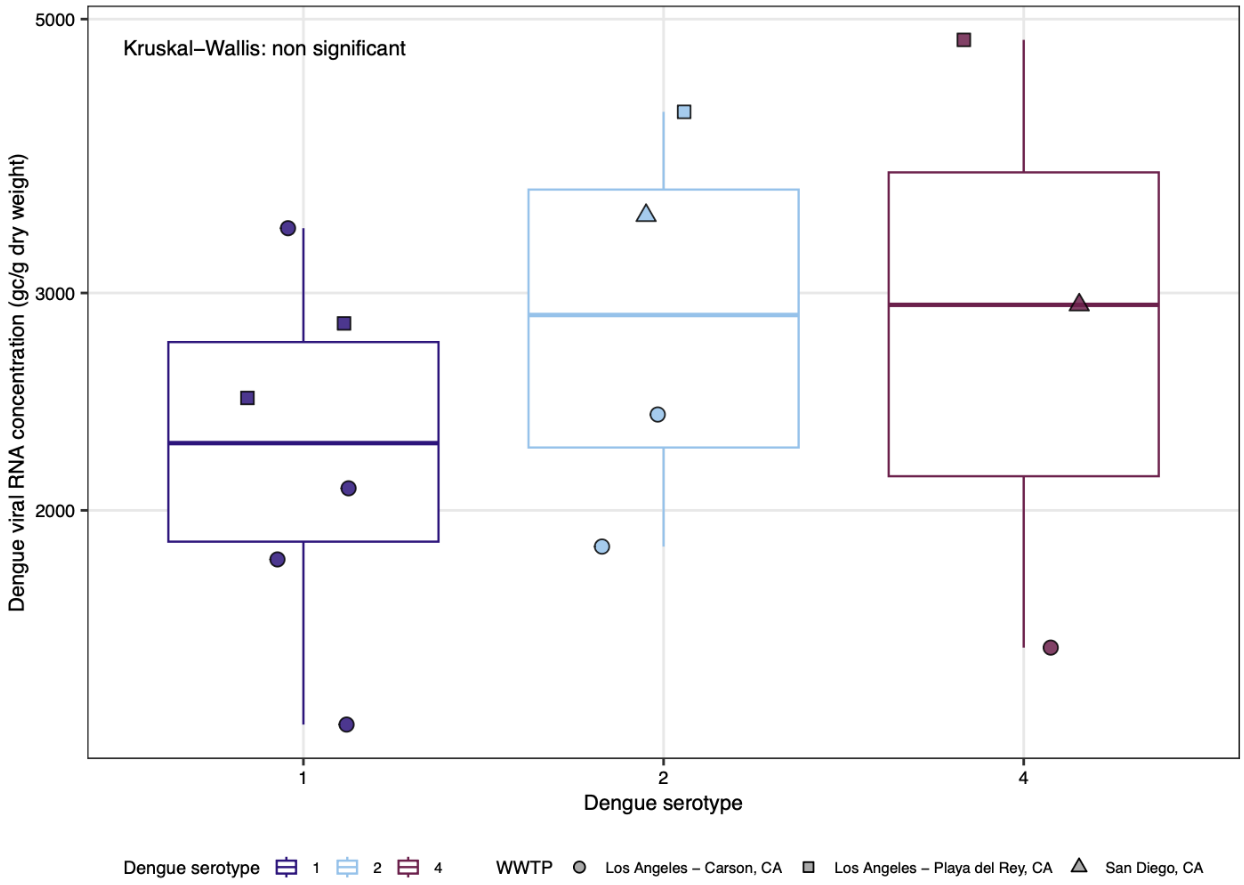


Supplementary Figure 1: Distribution of dengue viral RNA concentrations by serotype across wastewater treatment plants (WWTPs). Boxplots show the distribution of dengue viral RNA concentrations (gc/g dry weight) in wastewater solids for each detected dengue virus serotype across all sampled WWTPs. Points represent individual wastewater samples, with symbols indicating WWTP location. Only samples with detectable concentrations are included. Boxes represent the interquartile range (IQR), horizontal lines indicate medians, and whiskers extend to 1.5×IQR. DENV-3 is not shown because no detections were observed during the study period.

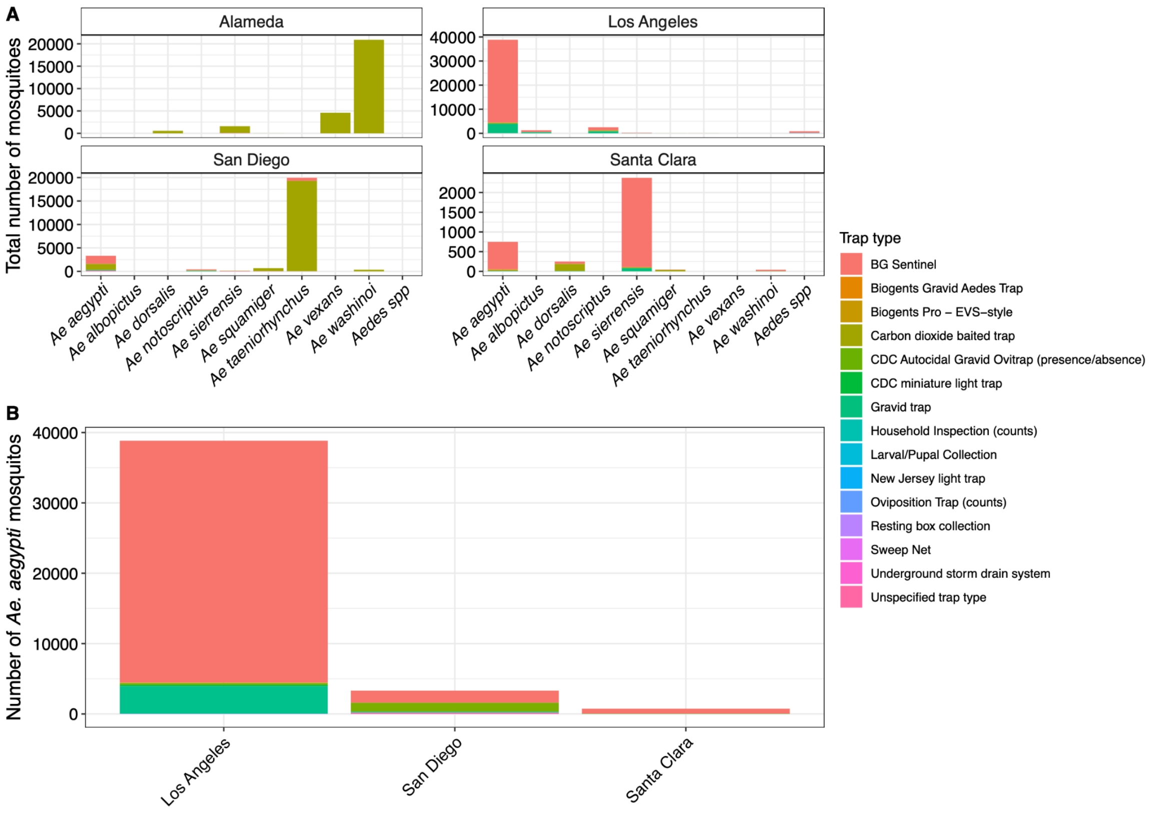


Supplementary Figure 2: Distribution of mosquito species and trap types across study counties. Stacked bar plots showing the total number of mosquitoes collected across four California counties (Alameda, Los Angeles, San Diego, and Santa Clara), stratified by trap type. Mosquito collection data were filtered to the wastewater sampling window of each county. Colors indicate different trap types used for collection. (A) Species-level distributions within each county. (B) Total *Aedes aegypti* mosquito counts by county and trap type.

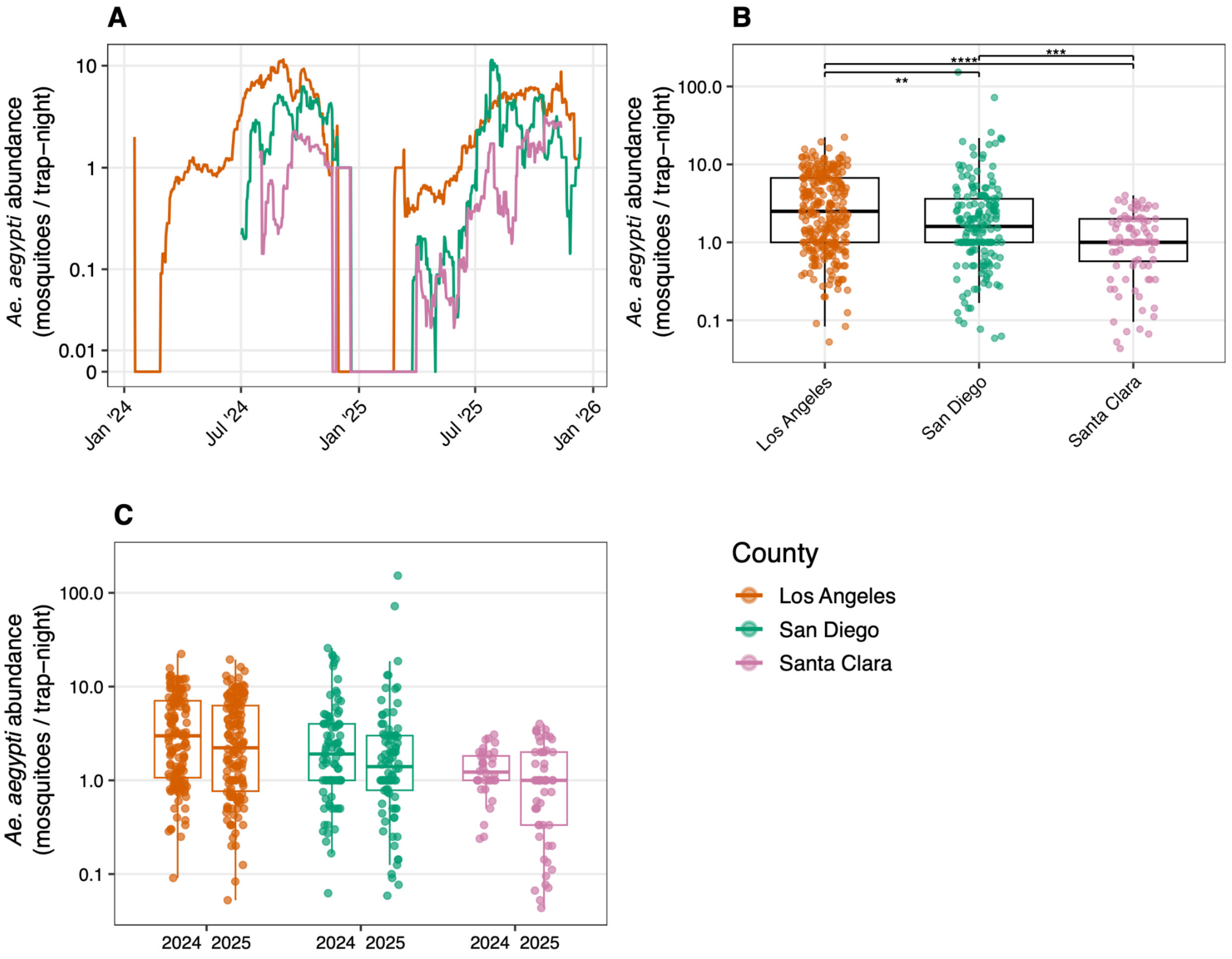


Supplementary Figure 3: Seasonal dynamics and distribution of *Aedes aegypti* mosquito abundance, standardized by trapping effort (mosquitoes per trap-night), across California counties during the wastewater sampling period. Mosquito surveillance data were restricted to the county-specific wastewater collection windows. Alameda County is not shown because no *Ae. aegypti* mosquitoes were detected during the wastewater sampling period corresponding to Oakland, CA. (A) Daily *Ae. aegypti* abundance (mosquitoes per trap-night) over time, shown as the centered 15-day rolling average for each county. Colors indicate county. (B) Distribution of daily *Ae. aegypti* abundance (mosquitoes per trap-night) across counties over the full wastewater collection window for each county. Boxplots summarize daily standardized mosquito abundance (median and interquartile range), with individual observations overlaid and colors indicating county. Horizontal brackets indicate significant pairwise differences between counties based on Dunn's test with Benjamini-Hochberg correction. (C) Distribution of daily *Ae. aegypti* abundance (mosquitoes per trap-night) by county and year. For each county, separate boxplots are shown for 2024 and 2025, with individual observations overlaid and colors indicating county. Horizontal brackets indicate significant differences between years within counties based on Wilcoxon rank-sum tests with Benjamini-Hochberg correction. Significance symbols indicate adjusted p-values as follows: * $p \leq 0.05$, ** $p \leq 0.01$, *** $p \leq 0.001$, and **** $p \leq 0.0001$.

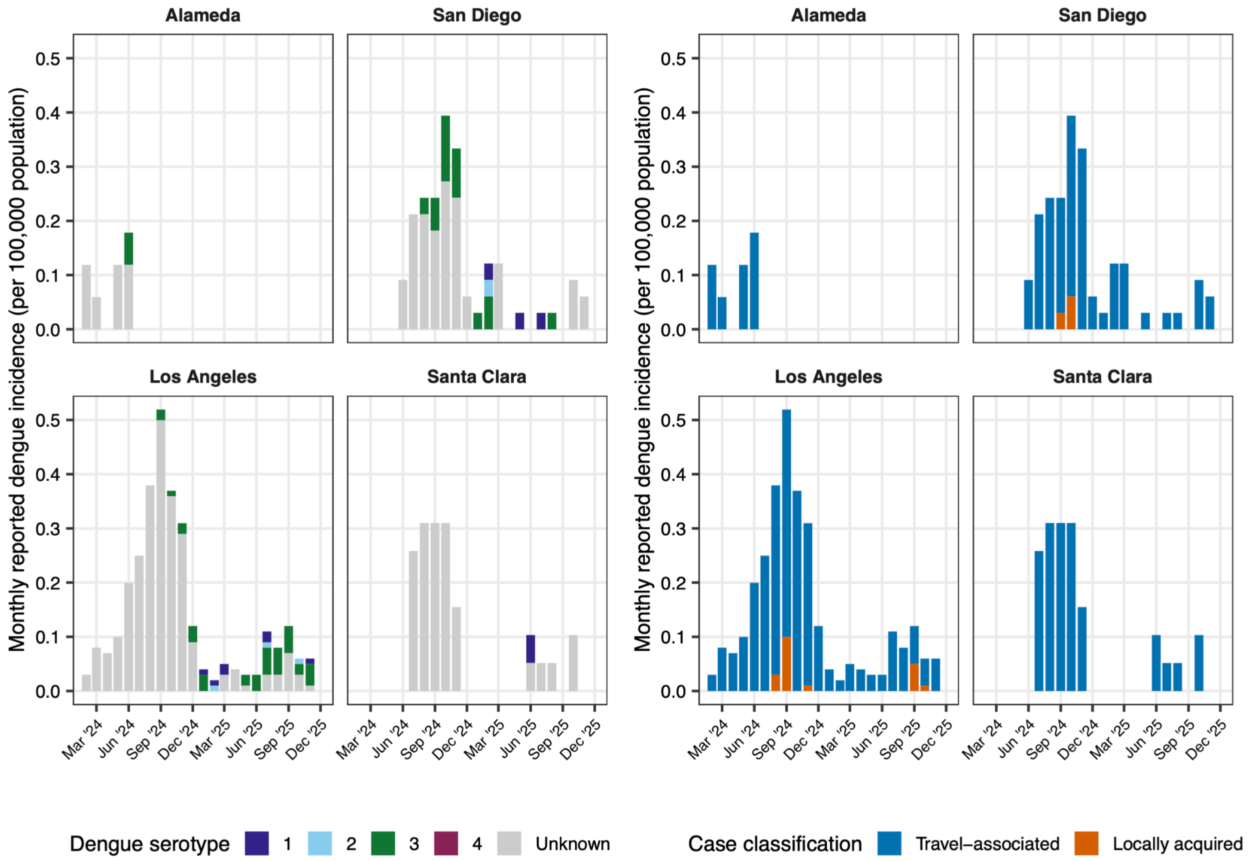


Supplementary Figure 4: Monthly aggregation of reported dengue incidence (per 100,000 population) in four California counties (Alameda, San Diego, Los Angeles, and Santa Clara) from January 2024 to December 2025, based on week of symptom onset. (A) Bars represent reported dengue incidence and are colored by reported dengue virus (DENV) serotype (DENV-1 to DENV-4). Grey bars indicate cases for which the infecting serotype was unknown. (B) The same incidence classified by travel status, colored by whether the case had a reported travel history prior to diagnosis (travel-associated) or no reported travel history (locally acquired).

### 2.2 Supplementary Tables

Supplementary Table 1: Primer and probe sequences used for serotype-specific detection of dengue virus (DENV-1 to DENV-4) by probe-based RT-ddPCR. Forward and reverse primers and hydrolysis probes (5′→3′) targeting conserved genomic regions of each dengue virus serotype are listed. Degenerate bases follow IUPAC nucleotide nomenclature. Probes were labeled with serotype-specific fluorophores (HEX for DENV-1, FAM for DENV-2, ROXN for DENV-3, ATTO590N for DENV-4) and dual-quenched (ZEN/3IABkFQ). For DENV-3, both standard and degenerate probe variants were evaluated to account for sequence variability.

| | | |
|---|---|---|
| DENV 1 | Forward | CAAAAGGAAGTCGYGCAATA |
| | Reverse | CTGAGTGAATTCTCTCTGCTRAAC |
| | Probe | /5hex/CATGTGGYT/zen/GGGAGCRCGC/3IABkFQ/ |

| | | |
|---|---|---|
| DENV 2 | Forward | CAGGCTATGGCACYGTCACGAT |
| | Reverse | CCATYTGCAGCARCACCATCTC |
| | Probe | /56-fam/CTCYCCRAG/zen/AACGGGCCTCGACTTCAA/3IABkFQ/ |
| DENV 3 | Forward | GGACTRGACACACGCACCCA |
| | Reverse | CATGTCTCTACCTTCTCGACTTGYCT |
| | Probe | /56-roxn/ACCTGGATG/zen/TCGGCTGAAGGAGCTTG/3IABkFQ/ |
| | Probe_degen | /56-roxn/ACCTGGATG/zen/TCGGCTGAAGGRGCTTG/3IABkFQ/ |
| DENV 4 | Forward | TTGTCCTAATGATGCTRGTCG |
| | Reverse | TCCACCYGAGACTCCTTCCA |
| | Probe | /5atto590n/TYCCTACYC/zen/CTACGCATCGCATTCCG/3IABkFQ/ |

Supplementary Table 2: In silico evaluation of primer and probe complementarity for dengue virus (DENV) RT-ddPCR assays. Publicly available DENV genomes were downloaded from NCBI and restricted to human-derived sequences with reported collection dates between July 1, 2023 and December 31, 2025. For DENV-3, results are shown separately for sequences evaluated with the original (DENV-3 original) and updated (DENV-3 degenerated) probe configurations. For each serotype, the table reports the percentage and number of sequences with perfect matches, primer mismatches only, probe mismatches, or failed screening. Fail indicates that no candidate amplicon met the screening thresholds of ≤2 mismatches in each primer and ≤1 mismatch in the probe. Percentages were calculated relative to the total number of sequences screened per serotype.

| **Serotype** | **Perfect match (0 mismatches)** | **Primer mismatch only** | **Probe mismatch** | **Fail** |
|---|---|---|---|---|
| DENV-1 | 41.3% (173/419) | 50.1% (210/419) | 5.3% (22/419) | 3.3% (14/419) |
| DENV-2 | 10% (85/846) | 7.7% (65/846) | 59.7% (505/846) | 22.6% (191/846) |
| DENV-3 degenerated | 83.5% (670/802) | 6.1% (49/802) | 10.1% (81/802) | 0.2% (2/802) |
| DENV-3 original | 16% (128/802) | 2.6% (21/802) | 78.6% (630/802) | 2.9% (23/802) |
| DENV-4 | 70.1% (61/87) | 9.2% (8/87) | 14.9% (13/87) | 5.7% (5/87) |

Supplementary Table 3: Full per-plate ddPCR quality-control results across the study period. Each row reports one control-target combination tested on a given weekly ddPCR plate. "week_start" and "week_end" give the first and last day of that week sample-processing period. "control" indicates the control type included on the plate, namely a no-template control (NTC) or a ddPCR positive control. "target" indicates the serotype-specific RT-ddPCR assay against which the control was run (DENV-1 to DENV-4). "positive_droplets" and "accepted_droplets" report the number of positive and total accepted droplets for that control-target reaction. "positive_call" indicates whether the reaction was classified as positive (TRUE) or negative (FALSE), using the same ≥3 positive-droplet threshold applied to wastewater samples. This table is provided in the Excel file containing the supplementary tables. This table is available in the supplementary_tables.xlsx file.

Supplementary Table 4: Summary statistics of dengue virus (DENV) RNA concentrations (gc/g dry weight) in wastewater samples with quantified detections, stratified by wastewater treatment plant (WWTP) and serotype. Concentrations were measured using serotype-specific RT-ddPCR assays targeting DENV-1 to DENV-4. The table reports the number of samples with detection (n) and descriptive statistics of the measured concentrations among detected samples only (mean ± SD, median with interquartile range (IQR), and minimum-maximum). The row “All WWTPs” summarizes concentrations pooled across all eight WWTPs. Values reported as “ND” (not detected) were excluded from concentration summaries. DENV-3 is not included because no detections were observed during the study period.

| WWTP | Serotype | Detected samples (n) | Mean ± SD | Median (IQR) | Min - Max |
|---|---|---|---|---|---|
| **All WWTPs** | DENV-1 | 6 | 2322.84 ± 732.06 | 2275.36 (1889.88 - 2742.50) | 1341.32 - 3385.39 |
| | DENV-2 | 4 | 2983.37 ± 1051.76 | 2929.13 (2261.11 - 3651.39) | 1869.48 - 4205.76 |
| | DENV-4 | 3 | 3097.53 ± 1636.95 | 2934.26 (2241.29 - 3872.12) | 1548.33 - 4809.99 |
| **Los Angeles - Carson, CA** | DENV-1 | 4 | 2158.98 ± 873.63 | 1954.61 (1704.19 - 2409.39) | 1341.32 - 3385.39 |
| | DENV-2 | 2 | 2130.57 ± 369.23 | 2130.57 (2000.03 - 2261.11) | 1869.48 - 2391.66 |
| | DENV-4 | 1 | 1548.33 ± NA | 1548.33 (1548.33 - 1548.33) | 1548.33 - 1548.33 |
| **Los Angeles - Playa del Rey, CA** | DENV-1 | 2 | 2650.55 ± 260.07 | 2650.55 (2558.61 - 2742.50) | 2466.66 - 2834.45 |
| | DENV-2 | 1 | 4205.76 ± NA | 4205.76 (4205.76 - 4205.76) | 4205.76 - 4205.76 |
| | DENV-4 | 1 | 4809.99 ± NA | 4809.99 (4809.99 - 4809.99) | 4809.99 - 4809.99 |
| **San Diego, CA** | DENV-2 | 1 | 3466.60 ± NA | 3466.60 (3466.60 - 3466.60) | 3466.60 - 3466.60 |
| | DENV-4 | 1 | 2934.26 ± NA | 2934.26 (2934.26 - 2934.26) | 2934.26 - 2934.26 |

Supplementary Table 5: Statistical comparison of detected dengue virus (DENV) RNA concentrations among serotypes pooled across all wastewater treatment plants (WWTPs). The global difference among serotypes was assessed using a Kruskal-Wallis. Pairwise differences were evaluated using Wilcoxon rank-sum tests with Benjamini-Hochberg adjustment. ‘ns’ indicates non significant. Comparisons with DENV-3 are not displayed because the serotype was not detected.

| **All serotypes - Kruskal-Wallis** | | p-value = 0.52 | | |
|---|---|---|---|---|
| **Comparison** | **Serotype 1** | **Serotype 2** | **BH-adjusted p-value** | **Significance** |
| Wilcoxon rank-sum | DENV-2 | DENV-1 | 0.78 | ns |
| Wilcoxon rank-sum | DENV-4 | DENV-1 | 0.78 | ns |
| Wilcoxon rank-sum | DENV-4 | DENV-2 | 1 | ns |

Supplementary Table 6: Statistical comparison of daily *Aedes aegypti* abundance among California counties. Differences among counties were evaluated using the Kruskal-Wallis rank-sum test, followed by pairwise Dunn's tests with Benjamini-Hochberg adjustment for multiple comparisons. Adjusted p-values are shown for pairwise comparisons.

| Test | Comparison | Statistic | Effect size | p-value | Adjusted p-value |
|---|---|---|---|---|---|
| Kruskal-Wallis | Across counties | 42.6 | 0.07 | <0.001 | NA |
| Dunn | Los Angeles vs San Diego | -3.21 | NA | 0.001 | 0.001 |
| Dunn | Los Angeles vs Santa Clara | -6.4 | NA | < 0.001 | < 0.001 |
| Dunn | San Diego vs Santa Clara | -3.55 | NA | < 0.001 | < 0.001 |

Supplementary Table 7: Summary statistics of *Aedes aegypti* mosquito abundance by county across the full study period and by calendar year (2024-2025).

| County | Period | Number of days | Mean abundance (mosquitoes/trap-night) | SD | Median abundance (mosquitoes/trap-night) | IQR | Minimum | Maximum | Total mosquitoes | Total trap-nights | Overall abundance (mosquitoes/trap-night) |
|---|---|---|---|---|---|---|---|---|---|---|---|
| Los Angeles | Overall | 707 | 1.6 | 3.2 | 0.0 | 1.4 | 0.0 | 22.3 | 38844 | 8392 | 4.6 |
| | 2024 | 357 | 1.7 | 3.3 | 0.0 | 1.5 | 0.0 | 22.3 | 20432 | 3720 | 5.5 |
| | 2025 | 350 | 1.5 | 3.1 | 0.0 | 1.4 | 0.0 | 19.4 | 18412 | 4672 | 3.9 |
| San Diego | Overall | 543 | 1.4 | 7.7 | 0.0 | 0.8 | 0.0 | 152.5 | 3318 | 1228 | 2.7 |

| | | | | | | | | | | | |
|---|---|---|---|---|---|---|---|---|---|---|---|
| | 2024 | 190 | 1.7 | 4.0 | 0.0 | 1.5 | 0.0 | 25.8 | 1666 | 540 | 3.1 |
| | 2025 | 353 | 1.2 | 9.1 | 0.0 | 0.1 | 0.0 | 152.5 | 1652 | 688 | 2.4 |
| Santa Clara | Overall | 484 | 0.2 | 0.7 | 0.0 | 0.0 | 0.0 | 4.0 | 749 | 803 | 0.9 |
| | 2024 | 161 | 0.3 | 0.7 | 0.0 | 0.0 | 0.0 | 3.1 | 331 | 266 | 1.2 |
| | 2025 | 323 | 0.2 | 0.7 | 0.0 | 0.0 | 0.0 | 4.0 | 418 | 537 | 0.8 |

Supplementary Table 8: Statistical comparison of daily *Aedes aegypti* abundance between calendar years within each California county. Differences between 2024 and 2025 were evaluated separately within each county using Wilcoxon rank-sum tests, with Benjamini-Hochberg adjustment for multiple comparisons across counties. W represents the Wilcoxon test statistic, and adjusted p-values are reported.

| **County** | **Comparison** | **W statistic** | **p-value** | **Adjusted p-value** |
|---|---|---|---|---|
| Los Angeles | 2024 vs 2025 | 11109 | 0.09 | 0.13 |
| San Diego | 2024 vs 2025 | 4319 | 0.14 | 0.14 |
| Santa Clara | 2024 vs 2025 | 1246 | 0.08 | 0.13 |

Supplementary Table 9: Summary statistics of weekly reported dengue incidence by county during the study period. Weekly case counts were aggregated by week of symptom onset and summed across dengue virus (DENV) serotypes, then divided by county population (2) and multiplied by 100,000 to give a weekly incidence value. Weeks without reported dengue cases were retained as zero-incidence weeks. Cumulative reported dengue incidence is the total number of reported cases over the whole study period divided by county population and multiplied by 100,000. The table reports county population, the total number of reported cases over the study period, the cumulative study-period incidence, summary statistics of weekly incidence (mean with SD, median with IQR, minimum, and maximum), the number and percentage of cases associated with each DENV serotype (DENV-1 to DENV-4) and cases with unknown serotype, the number and percentage of cases classified as travel-associated versus locally acquired, and the number and percentage of travel-associated and locally acquired cases broken down by year. Serotype and overall travel-history percentages are relative to the total number of reported cases within each county; year-specific travel-history percentages are relative to the total cases within each county and year.

| **County** | **County population** | **Total reported dengue cases** | **Cumulative dengue incidence (per 100,000 population)** | **Mean weekly reported dengue incidence ± (SD)** | **Median weekly reported dengue incidence (IQR)** | **Minimum weekly reported dengue incidence** | **Maximum weekly reported dengue incidence** | **DENV-1 reported cases, n (%)** | **DENV-2 reported cases, n (%)** | **DENV-3 reported cases, n (%)** | **DENV-4 reported cases, n (%)** | **Unknown serotype reported cases, n (%)** | **Travel-associated cases, n (%)** | **Locally acquired cases, n (%)** | **2023 Travel-associated cases, n (%)** | **2023 Locally acquired cases, n (%)** | **2024 Travel-associated cases, n (%)** | **2024 Locally acquired cases, n (%)** |
|---|---|---|---|---|---|---|---|---|---|---|---|---|---|---|---|---|---|---|
| Los Angeles | 10,014,009 | 320 | 3.2 | 0.03 ± 0.03 | 0.02 (0.03) | 0 | 0.15 | 7 (2.2%) | 3 (0.9%) | 37 (11.6%) | 0 (0.0%) | 273 (85.3%) | 300 (93.8%) | 20 (6.2%) | 3 (100.0%) | 0 (0.0%) | 238 (94.4%) | 14 (5.6%) |
| San Diego | 3,298,634 | 69 | 2.09 | 0.02 ± 0.03 | 0.00 (0.03) | 0 | 0.15 | 3 (4.3%) | 1 (1.4%) | 14 (20.3%) | 0 (0.0%) | 51 (73.9%) | 66 (95.7%) | 3 (4.3%) | 0 (0.0%) | 0 (0.0%) | 49 (94.2%) | 3 (5.8%) |

| | | | | | | | | | | | | | | | | | | |
|---|---|---|---|---|---|---|---|---|---|---|---|---|---|---|---|---|---|---|
| Santa Clara | 1,936,259 | 32 | 1.65 | 0.02 ± 0.04 | 0.00 (0.00) | 0 | 0.21 | 1 (3.1%) | 0 (0.0%) | 0 (0.0%) | 0 (0.0%) | 31 (96.9%) | 32 (100.0%) | 0 (0.0%) | 0 (0.0%) | 0 (0.0%) | 26 (100.0%) | 0 (0.0%) |
| Alameda | 1,682,353 | 13 | 0.77 | 0.01 ± 0.02 | 0.00 (0.00) | 0 | 0.12 | 0 (0.0%) | 0 (0.0%) | 1 (7.7%) | 0 (0.0%) | 12 (92.3%) | 13 (100.0%) | 0 (0.0%) | 1 (100.0%) | 0 (0.0%) | 12 (100.0%) | 0 (0.0%) |

Supplementary Table 10: County-level summary of wastewater sampling, DENV detection, and reported dengue cases and incidence across the four California counties. For each county, wastewater results are pooled across all included WWTPs; reported case counts are restricted to the wastewater sampling window of that county (first to last sample date) for comparability. Cumulative dengue incidence was calculated as the total number of reported cases within that window divided by the population of the county retrieved from the (2), and multiplied by 100,000. This table underlies panel B of Figure 3 and Supplementary Table 11.

| **County** | **WWTPs included** | **Wastewater samples** | **DENV-positive samples** | **Wastewater detection frequency (%)** | **Total reported dengue cases** | **Cumulative dengue incidence (per 100,000 population)** |
|---|---|---|---|---|---|---|
| Alameda | 1 | 81 | 0 | 0 | 13 | 0.8 |
| Los Angeles | 2 | 548 | 11 | 2 | 320 | 3.2 |
| San Diego | 1 | 212 | 2 | 0.9 | 69 | 2.1 |
| Santa Clara | 1 | 228 | 0 | 0 | 32 | 1.7 |

Supplementary Table 11: Rank correlations for the sample-/week-level comparisons in Figure 3. (A) Wastewater DENV concentration vs. same-week reported dengue incidence (per 100,000 population), one point per wastewater sample. (B) Wastewater detection frequency vs. cumulative reported dengue incidence (per 100,000 population) over the whole wastewater sampling window, one point per county (n = 4; interpret descriptively). (C) Weekly *Ae. aegypti* abundance vs. weekly reported dengue incidence (per 100,000 population), one point per county-week (all cases). (D) Same as (C), restricted to locally acquired cases only.

| **Panel** | **Comparison** | **Spearman ρ** | **p-value** | **n** |
|---|---|---|---|---|
| A | Wastewater DENV concentration vs weekly dengue incidence | 0.04 | 0.15 | 1069 |
| B | Wastewater DENV detection frequency vs cumulative dengue incidence | 0.95 | NA | 4 |
| C | Weekly mosquito abundance vs weekly dengue incidence | 0.39 | < 0.001 | 246 |
| D | Weekly mosquito abundance vs weekly dengue incidence of locally-transmitted cases | 0.45 | < 0.001 | 124 |
